# Propagation-based phase-contrast breast computed tomography: a visual grading assessment of the performance of photon-counting and flat-panel X-ray detectors

**DOI:** 10.1101/2022.11.01.22281633

**Authors:** Nicola Giannotti, Seyedamir Tavakoli Taba, Timur Gureyev, Sarah Lewis, Luca Brombal, Renata Longo, Sandro Donato, Giuliana Tromba, Lucia Arana Pena, Daniel Hausermann, Chris Hall, Anton Maksimenko, Benedicta Arhatari, Yakov Nesterets, Patrick Brennan

## Abstract

**Rationale and objectives:** Breast cancer represents the leading cause of death from cancer in women worldwide. Early detection of breast tumours improves the prognosis and survival rate. Propagation-based phase-contrast computed tomography (PB-CT) is a technique that uses refraction and absorption of the X-ray to produce images for clinical applications. This study compared the performance of photon-counting and flat-panel X-ray detectors in PB-CT breast imaging using synchrotron radiation.

**Materials and methods:** Mastectomy specimens underwent PB-CT imaging using the Hamamatsu C10900D Flat Panel and PIXIRAD-8 CdTe single-photon-counting detectors. PB-CT images generated at different imaging conditions were compared to absorption-based CT (AB-CT) reference images acquired with the same detectors to investigate the image quality improvement delivered by PB-CT relative to AB-CT. The image quality of the different image sets was assessed by eleven readers in a visual grading characteristics (VGC) study.

**Results:** The intraclass correlation coefficient showed a moderate/good interobserver agreement for the image set analysed (ICC = 0.626, p = <0.001). The area under the curve showed that the image quality improvement in PB-CT images obtained by the PIXIRAD-8 CdTe single-photon-counting detector were consistently higher than the one for flat-panel Hamamatsu detector. The level of improvement in image quality was more substantial at lower radiation doses.

**Conclusion:** In this study, the PIXIRAD-8 photon-counting detector was associated with higher image quality scores at all tested radiation dose levels, which was likely a result of the combined effect of the absence of dark current noise and better spatial resolution, compared to the flat-panel detector.

## Introduction

Breast cancer represents the leading cause of death from cancer in women worldwide^1^. Every year over 2 million breast cancers are diagnosed across the globe, and in 2018 alone GLOBOCAN estimated over 600,000 deaths related to breast cancer^2^. The lifetime risk of a woman developing breast cancer has been established as one in eight^3^, and it is commonly accepted that early detection of breast tumours increases available treatment options and improves the prognosis of patients and survival rate^4^.

Today, digital mammography (MG) and digital breast tomosynthesis (DBT) represent the two gold-standard imaging techniques used in breast cancer screening programs to detect early neoplastic lesions; nevertheless, several limitations related to the current technology and breast parenchymal components affect their diagnostic efficacy^5, 6^. Recent data showed that the false-negative rate in MG screening investigations is about 30% in women presenting with non-dense breast parenchyma^7^; however, the false-negative MG screening rate may increase up to 50% in the sub-group of women presenting with denser breast parenchyma^8^. The false-positive rate of breast screening programs is also not optimal, with a 7 to 12 % of screened women being unnecessarily re-called for further diagnostic investigations due to suspicious lesions detected on their initial scans^9^. In real life, this translates into increased cost, patient discomfort and longer patient waiting lists^10^.

Although different technologies are involved in the production of MG and DBT breast images, both systems use the basic property of X-ray attenuation through the matter to differentiate normal breast parenchyma from neoplastic lesions. However, in breast imaging the information received from soft tissue contrast differences may not be sufficient to identify neoplastic lesions due to the small tissue density differences existing between pathological and non-pathological tissues^11,19,20^. Additionally, not being fully-3D techniques, both MG and DBT are affected by anatomical noise potentially hampering the diagnosis. For this reason, newer technological advancements are needed to enhance lesion visualisation, reduce the frequency of false-negative, reduce patient recalls and finally improve patient risk-stratification.

Propagation-based phase-contrast (PB-CT) is emerging as a promising diagnostic technique capable of providing fully-3D images with high contrast-to-noise ratio (i.e. high visibility) at clinically compatible radiation dose levels^21^. Particularly, in addition to attenuation, PB-CT is able to detect phase-contrast effects due to the phase shifts occurring to X-ray waves when travelling through matter^12^ that, in soft-tissues and energies of radiological interest (10-100 keV), are larger than conventional relative attenuation contrast^13,14^

Due to the X-ray coherence requirements of propagation-based imaging, synchrotron facilities are currently being predominantly used in the development of PB-CT breast imaging clinical studies and, prospectively, to investigate the optimal imaging conditions that may be adopted in future to produce diagnostic images in more compact clinical systems. PB-CT has not yet been validated as an alternative to MG and DBT for screening and diagnostic purposes, and scanning parameters and hardware necessary to produce the highest quality diagnostic images are still being investigated in preparation for the clinical trials that are foreseen in the next 1-2 years^22,23^. Previous research has determined the best imaging conditions for the breast PB-CT at synchrotron facilities (with phantoms, formalin-fixed specimens and fresh human tissues) in terms of X-ray energy, sample-to-detector distance and algorithms used for phase-retrieval and reconstruction^24, 25,26,27,27^.

In this context, the choice of the imaging detector plays a crucial role in the final image quality. Since the advent of digital radiography, indirect conversion detectors have been the most widely used devices, being nowadays a mature technology. In recent years, thanks to the developments in semiconductors manufacturing, direct conversion photon-counting detectors with relatively high atomic number sensors are becoming a viable alternative to conventional devices. Thanks to the electronics embedded in each pixel, photon counters are able to discriminate every individual photon enabling a full electronic noise rejection. High-Z sensors, such as CdTe, ensure a high absorption efficiency being suitable for applications where strict limitations in terms of dose are present. Moreover, thanks to the direct conversion mechanism of X-ray photons into electrical charge, these devices usually offer a pixel-size limited spatial resolution, whereas conventional detectors are usually limited by the blurring due to the conversion of deposited X-ray energy into visible light^28^. The latter feature is of great importance for PB-CT, as a high response sharpness ensures optimal detection of phase-contrast effects (i.e., edge-enhancement)^26,29,30^.

In this framework, the aim of this study was to compare the performance of photon-counting and flat- panel X-ray detectors used in PB-CT breast cancer imaging, to assess the difference between these two technologies towards clinical applications. Specifically, absorption-based images acquired at a virtually null propagation distance and a reference radiation dose level, have been compared through a visual grading analysis study to phase-contrast images acquired at different propagation distances and doses with two detector technologies. The study is based on images of excised breast specimens scanned in the two participating centres (SYRMEP beamline of Elettra Synchrotron and IMBL at the Australian Synchrotron) working at the clinical translation of PB-CT.

## Methods

Ethical approval was received from the Human Research Ethics Committee of the University of Sydney (project number: CF15/3138-2015001340) for the scans performed at the imaging and medical beamline (IMBL). The scans at SYnchrotron Radiation for MEdical Physics (SYREMP) (Trieste, Italy) were performed according to the Directive 2004/23/EC of the European Parliament and of the Council of March 31^st^ 2004, on setting standards of quality and safety for the donation, procurement, testing, processing, preservation, storage, and distribution of human tissues and cells and in the framework of the operative protocol of the Institutional Review Board of the Breast Unit of the Trieste University Hospital (“PDTA Neoplasia mammaria”, ethical approval received on the 11th December 2019 by ASUGI-Azienda Sanitaria Universitaria Giuliano Isontina, Italy). Written consent was obtained from patients. No fixation and preservation were applied to the samples as all specimens were scanned within a few hours of the surgical excision. Six mastectomy specimens were scanned at the IMBL and three mastectomy pecimens were scanned at the SYREMP beamline for this study. Similar scanning protocol settings were adopted by the two participating centres for the acquisition of both absorption-based CT (reference images) and PB-CT images (Table 1).

**Table 1.** Scanning settings used for mastectomy PB-CT in the two participating centres.

|  | **SYRMEP beamline** | **IMBL** |
| --- | --- | --- |
| **Detector type and pixel size** | PIXIRAD-8 CdTe single-photon-counting (60 $\mu\text{m}$) | Hamamatsu C10900D Flat Panel Sensor (100 $\mu\text{m}$) |
| **Slice thickness used in the VGA study** | 1 mm | 1 mm |
| **Slice imaging plane** | Coronal | Coronal |
| **Scanning energy (keV)** | 32 | 32 |
| **Short sample-to-detector distance and MGD** | 3.7 m ($R'= 3.2$ m)<br>@ 5 mGy, 2 mGy and 1 mGy | N/A |
| **Long sample-to-detector distance and MGD** | 9.6 m ($R'= 6.7$ m)<br>@ 5 mGy, 2 mGy and 1 mGy | 6 m ($R'= 5.7$ m)<br>@ 4 mGy and 2 mGy |
| **TIE-Hom phase retrieval** | Full | Half |
| **Reference image (absorption-based CT)** | 32 keV, 0.2 m, 5 mGy | 32 keV, 0.2 m, 4 mGy |

### Propagation-based phase-contrast images

IMBL and SYRMEP beamlines are independently optimizing their imaging setups to ensure the highest achievable image quality for the clinical application. In this context, rather than matching the experimental conditions, this study was carried out at the optimal, but different, configurations of each setup. At the Australian IMBL, a near-parallel X-ray beam with cross-sectional area of approximately 120 mm (horizontal) x 30 mm (vertical) at the distance of 134 m from the source, energy of 32 keV, and an energy resolution (DE/E) of about 10^−3^ was used. The X-ray detector was a Hamamatsu C10900D Flat Panel Sensor (Hamamatsu Photonics, Hamamatsu, Japan), with a pixel size of 100 um x 100 um, field of view of 1248 × 1248 pixels, that was operated at a frame rate of 17 fps. The measured spatial resolution of Hamamatsu C10900D detector was approximately 1.5 times the pixel size (∼150 um). Each mastectomy sample was placed in a thin walled (1 mm) cylindrical plastic container with the nipple positioned on the top. The surgical sutures and clips on the mastectomy specimens were used to orientate the breast (resembling a coronal view) before positioning the container on a rotation stage for CT scans^15^. PB-CT scans were collected at the sample-to-detector distance of 6 m (maximum achievable distance at IMBL). Considering the effect of magnification resulting from long source-to-sample distance of 134 m, the effective propagation distance for these scans was R’= 5.7 m^26,30^. PB-CT scans were collected at two different radiation doses: 4 mGy mean glandular dose (MGD) (referred to as “standard” dose) and 2 mGy MGD (referred to as “low” dose). For each scan at 4 mGy, 2400 projections with 0.075° angular steps and, for 2 mGy scans, 1200 projections with 0.15° angular steps were collected over the 180° rotation range of continuous data acquisition. Moreover, dark-current images (for removing the detector dark-current contribution) and flat-field images (for correcting uneven illumination) were collected during each acquisition. To compensate for the possible temporal variation of the incident beam intensity and dark current instabilities, half of the dark-current and flat-field images were collected before the sample scan and the other half collected after the sample scan.

The SYRMEP beamline features a laminar X-ray beam with a cross-sectional area of 140 mm x 3.5 mm at the sample position, that is located at 22.5 m from the X-ray source^31^. The detector was positioned at two different propagation distances of 9.6 m (maximum achievable distance at SYRMEP) and 3.7 m (the distance foreseen for clinical applications), resulting in effective propagation distances of 6.7 m and 3.2 m when taking into account the magnification effect. As for IMBL measurements, the X-ray energy of 32 keV was used for scanning the samples. The detector used in the study was a photon-counting device, Pixirad-8 (Pixirad imaging, Pisa, Italy), featuring a 650 µm thick CdTe sensor and a pixel pitch of 60 <u>µm</u>^32,33^. The detector was operated in “dead time free” mode with an energy threshold of 3 keV. Scans were collected at the maximum detector frame rate of 30 fps and consisted of 1200 projections acquired over 180 degrees in steps of 0.125 degrees, while the rotator was set to a constant speed of 4.5 degrees/second. Background corrections were applied by collecting flat-field images for each scan and, prior to image processing, detector-specific corrections were applied through a dedicated pre-processing procedure^34^. Similar to IMBL, the mean glandular dose was estimated starting from air-kerma measurements performed with a calibrated ionization chamber by applying conversion factors derived from a dedicated Monte-Carlo simulation^35,36^. Images at 3.7 m (“short” propagation distance) were obtained at three different MGD levels, namely 5 mGy (“standard” dose), 2 mGy MGD (“low” dose) and 1 mGy MGD (“very low” dose). At 9.6 m (“long” propagation distance) the X-ray flux was slightly increased to compensate for air attenuation, resulting in MGD values of 7 mGy (“standard”), 3 mGy (“low”) and 1.5 mGy (“very low” dose). It is worth mentioning that in a clinical setting the air attenuation can be virtually removed by positioning a vacuum pipe between sample (i.e., patient) and detector, hence bringing to a dose reduction to the patient, corresponding to 5 mGy, 2. mGy and 1 mGy, respectively, while keeping the same X-ray fluence at the detector.

After image acquisition and background/detector-specific corrections, projection images were processed through the Transport-of-Intensity Equation-based phase-retrieval filter (TIE-Hom)^38^. Considering previous optimization studies, the filter parameter γ was set to 870 (referred to as “Full” phase-retrieval) for the SYRMEP measurements, while it was set to 435 for IMBL measurements (referred to as “Half” phase-retrieval)^39,40^.

### Absorption-based CT images

Absorption-based CT (AB-CT) images were acquired at both centres using the smallest achievable sample-to-detector distance (0.2m) representing conventional computed tomography (CT) and the X-ray energy of 32 keV. AB-CT data were acquired and used in the subsequent analysis as reference images to compare the image quality of PB-CT images obtained using different detectors with reference to the AB-CT images. AB-CT scans were carried out using “standard” dose only (4 mGy MGD at IMBL and 5 mGy MGD at SYRMEP).

### Image assessment

Both PB-CT images and reference AB-CT images were reconstructed and displayed on coronal plane. PB-CT image quality of the data obtained at the two participating centres was subjectively assessed and compared to the relative AB-CT reference image by eleven imaging experts with at least 10 years of experience in medical imaging research and image quality evaluation in visual grading characteristics (VGC) studies. Image assessment was performed using dedicated workstations for breast imaging reporting. Each assessor was asked to compare the quality of the PB-CT images against “reference” AB-CT images of the same sample.

The image attributes were graded using a scale from -2 to +2, and were defined as follow:

- Perceptible contrast: difference between low and high radiolucency in various soft tissue regions.
- Lesion sharpness: clarity of definition of lesions and spiculations.
- Tissue interfaces: clarity of visualisation of interfaces between fatty and fibroglandular tissues.
- Calcification visibility: sharpness of micro-calcifications (if any).
- Image noise: presence of quantum mottle in the image.
- Artefacts: evidence of any other technical artefacts such as rings or distortions.

### Statistical analysis

The interobserver agreement was quantified using intraclass correlation coefficient test (ICC). The image quality was analysed using visual grading characteristic (VGC) analysis (VGC Analyzer software v1.0.2). The rating scores were treated as ordinal and there were no assumptions about the distribution of the data. For each image criterion, the cumulative distributions of the rating data for the test images were plotted against the reference images, resulting in a curve for which the area under the curve (AUCVGC) could measure the difference in the image quality of the two images sets. When interpreting the VGC analysis results, an AUCVGC of 0.5 indicates the equivalence of the two image sets, 0 ≤ AUCVGC < 0.5 indicates that the quality of the test images was lower than that of the reference images, and 0.5 < AUCVGC ≤ 1 indicates that the quality of the test images was higher than that of the reference images.

## Results

The ICC test showed a moderate/good interobserver agreement (ICC = 0.626, p = < 0.001). When the AB-CT images were used as a reference to compare the improvement in image quality of PB-CT images at different scanning conditions, the PIXIRAD-8 CdTe single-photon-counting detector (Figure 1) operating at the SYRMEP facility recorded significantly higher scores than the C10900D Flat Panel Detector (Figure 2). The PB-CT images obtained at IMBL using the Hamamatsu C10900D Flat Panel Sensor at the standard dose were of significantly higher image quality than standard dose AB_CT images (AUC = 0.803, p < 0.01); however, the corresponding low dose PB-CT images were not significantly different from the standard dose AB-CT images (Table 2).

**Figure 1:**
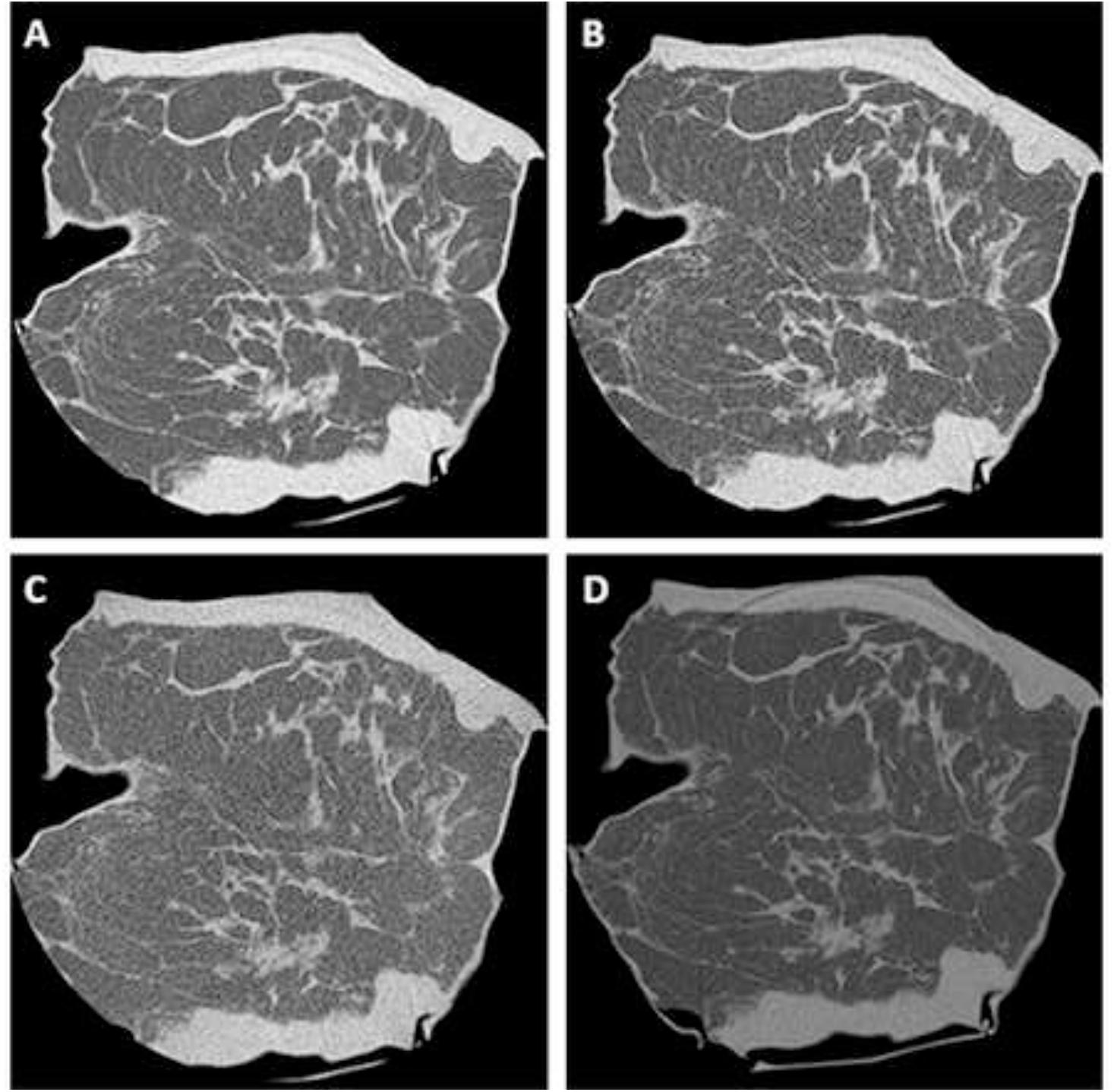
**A mastectomy sample scanned at the the Elettra Synchrotron (Trieste, Italy) at various imaging conditions. A) PB-CT sample-to-detector distance (9.6 m), dose 7 mGy; B) PB-CT sample-to-detector distance (9.6 m), dose 3 mGy; C) PB-CT sample-to-detector distance (9.6 m), dose 1.5 mGy; D) AB-CT sample to detector distance (0.2 m), dose 5 mGy (ref. image)**

**Figure 2:**
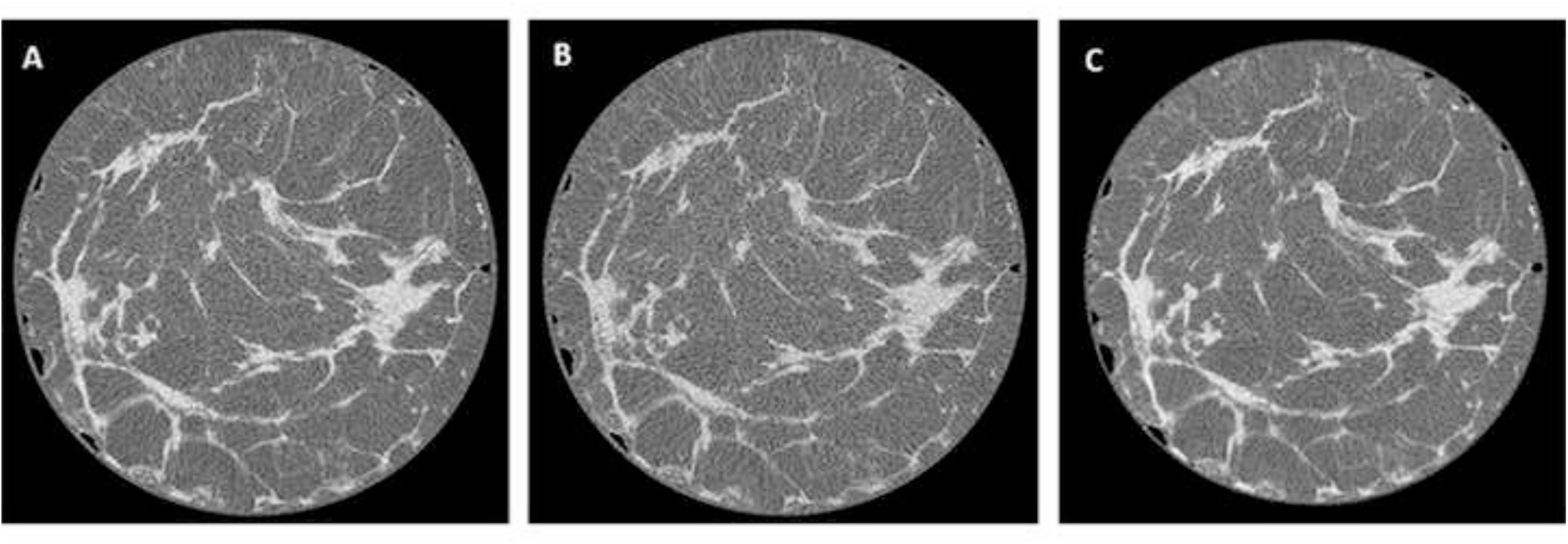
**A mastectomy sample scanned at the Australian Synchrotron at 32 keV and various imaging conditions. A) PB-CT sample-to-detector distance (6 m), dose 4 mGy; B) PB-CT sample-to-detector distance (6 m), dose 2 mGy; C) AB-CT sample to detector distance (0.2 m), dose 4 mGy (ref. image)**.

**Table 2.** SYRMEP vs Elettra image assessment scores – AB-CT images used as references.

| Beamline | Imaging condition | VGC |  |
| --- | --- | --- | --- |
|  |  | AUC | p-value |
| **SYRMEP**<br>PIXIRAD-8 CdTe single-photon-counting detector | 3.7m_1mGy | 0.000 | 0.263 |
|  | 3.7m_1mGy |  |  |
|  | 3.7m_1mGy |  |  |
|  | 3.7m_2mGy | 0.440 | 0.748 |
|  | 3.7m_2mGy |  |  |
|  | 3.7m_2mGy |  |  |
|  | 3.7m_5mGy | **0.970** | <0.001 |
|  | 3.7m_5mGy |  |  |
|  | 3.7m_5mGy |  |  |
|  | 9m_1.5mGy | 0.257 | <0.001 |
|  | 9m_1.5mGy |  |  |
|  | 9m_1.5mGy |  |  |
|  | 9m_3mGy | **0.955** | <0.001 |
|  | 9m_3mGy |  |  |
|  | 9m_3mGy |  |  |
|  | 9m_7mGy | **0.985** | <0.001 |
|  | 9m_7mGy |  |  |
|  | 9m_7mGy |  |  |
| **IMBL**<br>C10900D Flat Panel Detector | 6m_2mGy | 0.403 | 0.227 |
|  | 6m_2mGy |  |  |
|  | 6m_2mGy |  |  |
|  | 6m_2mGy |  |  |
|  | 6m_2mGy |  |  |
|  | 6m_2mGy |  |  |
|  | 6m_4mGy | **0.803** | <0.001 |
|  | 6m_4mGy |  |  |
|  | 6m_4mGy |  |  |
|  | 6m_4mGy |  |  |
|  | 6m_4mGy |  |  |
|  | 6m_4mGy |  |  |

The PB-CT images obtained at SYRMEP using the photon counting detector and long sample-to-detector distance at the standard dose and low dose were significantly superior to the standard dose AB-CT images (AUC = 0.985, p < 0.01 and AUC = 0.955, p < 0.01, respectively), showing that the improvement in image quality was much higher for the photon counting detector compared to the flat panel detector. Interestingly, at short sample-to-detector distance using photon counting detector, the standard dose PB-CT images were of higher quality than the standard dose AB-CT images (AUC = 0.970, p < 0.01), which denotes the improvement in image quality was higher than the one for the long sample-to-detector distance using the flat panel detector. Very low dose PB-CT scans were not superior to standard dose AB-CT images at any of the imaging conditions.

## Discussion

To our knowledge, this is the first study in assessing PB-CT image quality that tested the performances between photon counting and flat panels using an observer study. Using AB-CT images as a reference, this study compared the improvement in image quality of the breast PB-CT acquired with reasonably similar imaging conditions using different detector technologies. Our results showed that PIXIRAD-8 CdTe single-photon-counting detector can improve the quality of PB-CT images relevant to AB-CT images at a higher level than the C10900D Flat Panel Detector, even at the reduced radiation doses or shorter propagation distance used in combination with the photon counting detector.

Several factors are responsible for the different performances associated with the two detectors compared in this study. Compared to conventional flat panel detectors, in the photon counting detectors each pixel has its own circuitry able to read out the signal generated by individual X-ray photons, allowing the discrimination of electronic noise^18^. Moreover, thanks to the direct conversion of X-rays into electrical charges, these devices deliver a pixel size limited point-spread function (PSF) width (60 µm in the case of PIXIRAD8). Conversely, spatial resolution in indirect conversion devices is typically limited by the blurring due to the conversion of X-rays into visible light within the scintillator. For the Hamamtsu flat panel detector considered in this study this translates to a PSF of approximately 150 µm wide with a pixel size of 100 µm. We believe that both factors play an important role on the outcome of the final image quality in PB-CT.

To validate PB-CT as a breast screening or diagnostic tool, the detectors used must be able to produce high quality images capable of providing clinicians detailed information about early-stage breast lesion progression that may not be visible on standard mammograms, while using the same or lower radiation doses to the patients. Previous studies demonstrated that photon-counting PB-CT outperform clinical cone-beam breast CT^40,41^. The results from this study show that using a photon counting detector can enable a significant reduction of the radiation dose or propagation distance in the imaging setup, which are both critical for clinical translation of the PB-CT technology. Most importantly, the results from this study showed that high quality diagnostic PB-CT images can be generated using the photon-counting detector by reducing the measured MGD by at least 50%. To bring this into the context, the standard range of radiation doses delivered with current cone beam breast CT (CBBCT) is 4 to 12.8 mGy (mean, 8.2 [SD, 1.4] mGy)^17^, and our study showed that high quality PB-CT images can be effectively generated using reduced MGD in the range of 2 mGy.

The main limitations of this study are related to the limited sample size analysed and scanning settings that were similar but not the same. We acknowledge that the two experimental setups involved in this study used different imaging conditions such as sample to detector and source to detector distances, which is unavoidable considering the different layout of the two beamlines. Moreover, as previously mentioned, it should be stressed that each beamline developed its imaging protocol independently and that, in both cases, acquisition was performed in the best available experimental conditions. Although our results showed that Pixirad photon-counting detector can achieve higher image quality using lower radiation dose, further research involving larger datasets obtained using the same imaging condition is needed to validate and quantify the effect of photon-counting detectors on the improved image quality in PB-CT.

## Conclusion

The results from this initial work show that photon counting detectors (similar to PIXIRAD-8 CdTe used here) may provide improved image quality in the breast PB-CT at significantly reduced radiation doses compared to flat panel detectors. The results are promising in the development of PB-CT as a breast imaging diagnostic technique in the near future.

## Data Availability

All data produced in the present work are contained in the manuscript

## Notes

### Competing Interest Statement

The authors have declared no competing interest.

### Funding Statement

The authors declare that they have no known competing financial interests or personal relationships that could have appeared to influence the work reported in this paper.

### Author Declarations

Ethical approval was received from the Human Research Ethics Committee of the University of Sydney (project number: CF15/3138-2015001340) for the scans performed at the imaging and medical beamline (IMBL). The scans at SYnchrotron Radiation for MEdical Physics (SYREMP) (Trieste, Italy) were performed according to the Directive 2004/23/EC of the European Parliament and of the Council of March 31st 2004, on setting standards of quality and safety for the donation, procurement, testing, processing, preservation, storage, and distribution of human tissues and cells and in the framework of the operative protocol of the Institutional Review Board of the Breast Unit of the Trieste University Hospital ("PDTA Neoplasia mammaria", ethical approval received on the 11th December 2019 by ASUGI-Azienda Sanitaria Universitaria Giuliano Isontina, Italy). Written consent was obtained from patients.

